# Al-Powered classification of Ovarian cancers Based on Histopathological lmages

**DOI:** 10.1101/2024.06.05.24308520

**Authors:** Haitham Kussaibi, Elaf Alibrahim, Eman Alamer, Ghada Alhaji, Shrooq Alshehab, Zahraa Shabib, Noor Alsafwani, Ritesh G. Meneses

## Abstract

1

**Background:** Ovarian cancer is the leading cause of gynecological cancer deaths due to late diagnosis and high recurrence rates. While histopathological analysis is the gold standard for diagnosis, artificial intelligence (AI) models have shown promise in accurately classifying ovarian cancer subtypes from histopathology images. Herein, we developed an AI pipeline for automated identification of epithelial ovarian cancer (EOC) subtypes based on histopathology images and evaluated its performance compared to the pathologists’ diagnosis.

**Methods:** A dataset of over 2 million image tiles from 82 whole slide images (WSIs) of the major EOC subtypes (clear cell, endometrioid, mucinous, serous) was curated from public and institutional sources. A convolutional neural network (ResNet50) was used to extract features which were then input to 2 classifiers (CNN, and LightGBM) to predict the cancer subtype.

**Results:** Both AI classifiers achieved patch-level accuracy (97-98%) on the test set. Furthermore, adding a class-weighted cross-entropy loss function to the pipeline showed better discriminative performance between the subtypes.

**Conclusion:** AI models trained on histopathology image data can accurately classify EOC subtypes, potentially assisting pathologists and reducing subjectivity in ovarian cancer diagnosis.

## 2 Introduction

Ovarian cancer (OC) is the 3rd most common gynecological cancer worldwide (1) and the 7th in the Kingdom of Saudi Arabia among females (Saudi Council) (2). Due to the late stage of diagnosis and a high chance of recurrence (70%) (3), OC outranks other gynecological cancers in the mortality rate. Despite all the advanced developments in medical technology in diagnosis and treatment, OC is still discovered in the late stages which ultimately affects the treatment options and prognosis. Clinically, ultrasound and tumor biomarkers are used as the conventional diagnostic tools to investigate suspected cases of OC (4). However, histopathological study is the cornerstone to confirm the diagnosis (5). According to the World Health Organization, ovarian cancers are classified histologically into epithelial, germ cell, and sex-cord stromal tumors, in which epithelial ovarian cancer (EOC) stands out with the highest incident rate (6). In 2018, the Global Cancer Observatory (GLOBOCAN) reported that EOC accounts for 4.3% of cancer mortality annually, making EOC the most lethal gynecological cancer worldwide (7). EOC is a heterogeneous disease, that consists of five histological subtypes: High-Grade Serous Carcinoma (HGSC), Low-Grade Serous Carcinoma (LGSC), Mucinous Carcinoma (MC), Endometrioid Carcinoma (EC), and Clear Cell Carcinoma (CCC) (8). Each subtype exhibits unique histological morphology, molecular biology, pathogenesis, and clinical behavior (6).

Nowadays, the medical industry shows great interest in Artificial Intelligence (AI) implementation in different fields. According to Aliza Becker, AI usage in the medical field could be within one or more of the following categories: 1-For research purposes targeting pathology and\or treatment efficacy, 2-To minimize possible complications, 3-To maximize patient care during on-going treatment or procedure, and 4-In evaluating risk disease and predicting prognosis or treatment efficacy (9). Deep learning (DL), a subtype of AI, may achieve human-level accuracy. DL could predict clinical biomarkers, gene expression patterns, survival outcomes, and pathogenic mutations from traditional histoimage (10). Convolutional neural network (CNN) is an artificial neural network, preferred in image recognition and classification. Residual network (ResNet) is a type of CNN that has wide applications in image recognition and classification in medical and non-medical trials. Multiple studies were conducted respecting AI recruitment in active patient care (11).

In this study, digital histopathological images of various types of EOC were collected to create a dataset for training and validation of the AI system. Afterwards, we aimed to develop an AI model that can automatically identify EOC subtypes and evaluate the model’s performance by comparing it with the pathologist’s diagnosis.

In 2022, Rahman et al. worked with ResNet50 to measure its efficiency in diagnosing breast cancer by reading and classifying mammographic images. ResNet50 scored 93% in accuracy, with less effort and more time effectiveness (11). Myszczynska et al. shed light on machine learning utilization in the diagnosis and treatment of neurodegenerative diseases (14). The AI models reviewed in the earlier study are capable of analyzing a variety of clinical parameters such as neuroimages, electroencephalograms, and even cognitive test performance; suggesting an era of the machine learning revolution (12). The application of AI is becoming a trend in pathology settings, particularly in diagnosing cancer (10). Dolezal et al. emphasized the significant role of deep learning in analyzing digitized histopathological slides as it alleviates the workload on pathologists, detects features that could be easily missed, and even staining (13). ResNet50 has been used in histopathology image detection, for example in breast cancer and lung cancer diagnosis with 92% and 97.49% accuracy, respectively (14). However, in Saudi Arabia, AI in diagnostic radiology is the most studied and therefore most advanced in the health intelligence field (15). Further studies are essential to fuel the AI utilization in histopathology to rise to the advancement of other health fields locally. Additionally, the use of AI in EOC histopathology is poorly studied and further research is crucial to successfully implement it in clinical practice.

Several studies have discussed the use of AI in the diagnosis, classification, and prognosis of ovarian cancer. In 2018, researchers utilized Alexnet, a deep convolutional neural network (DCNN), to classify EOC into four subtypes serous, mucinous, endometrioid, and clear cell based on cytology (16). The study involved 87 Hematoxylin-Eosin (H&E) stained tissue samples which turned into a final of 81312 images, including augmented images. The DCNN consists of five conventional layers, three max-pooling layers, and two full reconnected layers. Aiming to improve network accuracy, this study used two different sets of input, one including the original slide set and the other with slide augmentation through different image rotation and edge sharpening filters. The classification accuracy was only 72.76% in total; with endometrioid being the lowest in accuracy (64.53%). Data augmentation proved its role in increasing total accuracy by 5.44%. Yet, the study has some limitations as the model incorrectly classified endometrioid as serous, mucinous as clear cell, and clear cell as mucinous with an error rate 15.11%, 12.64%, and 11.39%, respectively (16).

A retrospective cohort study was conducted to classify samples into either borderline ovarian tumor (SBOT) or high-grade serous ovarian cancer (HGSOC) (19). The study included 30 cases from the institutional pathology system database, half of each type. Researchers used a Support Vector Machine (SVM) classifier as an AI model to sort the sample based on 41 cellular features. By using Groovy scripts and QuPath, pathologists annotated the slides manually into stromal or tumor cells. Final results revealed accuracy of 86.4%-89.1% and 85.4%-90.8% for HGSOC and SBOT, respectively. Moreover, the overall SVM accuracy reached up to 90.5%-90.7%. One of the major features that played a crucial role in differentiation is Eosin OD staining intensity. Despite the high accuracy rate achieved, it is still strenuous for AI models to approximate human capacity. Multiple reasons for misclassification were observed; such as histological artifact and under/over cellular segmentation (17).

Two studies were conducted using a convolutional neural network (CNN) mainly to predict HGSOC response to platinum-based therapy and the prognosis accordingly (20,21). Liu et al. aimed to identify the morphological features that can enhance the decision-making towards a treatment plan and survival rate for patients (20). Researchers utilized 248 samples from The Cancer Genome Atlas (TCGA); 208 samples were used to train inception V3, and the remaining 40 were used for testing. To limit model bias, specialized pathologists selected regions of interest (ROI) from original slides and cropped them to tiles to fit into the input size, and these tiles were tagged as resistant or sensitive to chemotherapy. Moreover, training was repeated 16 times with parameter adjustments to ensure model generalizability. Another measure was used to size up and down the number of resistance or sensitive cases to assess if it would affect the model accuracy. Eventually, 85% of patients were categorized correctly, with a specificity of 90% and sensitivity of 73%. Furthermore, researchers stated that age, grade, and stage were not significantly associated with chemotherapy response. The main weakness of this study is the online data source, small validation, sample size, limited generalizability as it depends on TCGA only, and the data used were mostly high-grade tumors which decrease the clinical heterogeneity (18). Whereas Laury et al. used samples of patients diagnosed and treated for high-grade extrauterine serous carcinoma at HUS Helsinki University Hospital between 1982 and 2013 (21). 205 whole slide images were used to train the CNN and 22 slides were used to test the model. The model was capable of identifying 18 out of 22 testing slides with a sensitivity of 73% and a specificity of 91%. The positive predictive value is 89%, with an overall accuracy of 82%. The main weakness of this study is that molecular testing results were not available for all samples as the test was not routinely done (19).

In a retrospective cohort study conducted in 2020, Tanabe et al. used the CNN model AlexNet to analyze serum glycopeptides of ovarian cancer patients via converting glycopeptides expression into 2-dimensional (2D) barcode (20). A sample of 97 patients diagnosed with early-stage EOC were collected, in which 60% used for training and the remaining 40% for validation. Two sets of barcodes were prepared with different alignments based on liquid chromatography elution time and principal component analysis, resulting in area under the curve (AUC): 0.881 and 0.851, respectively. CA125 and HE4 information were added into the barcode to further enhance the diagnostic performance by changing the color (multicolored model). This results in increased diagnostics accuracy to 95%. Nonetheless, an Area Under the Receiver Operating Characteristic curve of almost 100% brings the model generalizability into question. Thus, extending training samples is a must to eliminate possible over-fitting (20). Another observational cohort study aimed to predict the mutation of BRCA 1/2 genes in ovarian cancer patients from H&E tissue using a CLAM-based approach (23). A sample of 664 ovarian cancer patients were involved, 464 utilized for the training process, and 132 for validation. The study showed disappointing results with low accuracy reaching 62.9% and 87.9% specificity with a 16.7% sensitivity, which suggests that phenotype may not be strongly related to genotype (21).

Other fields of research regarding AI and ovarian histopathological slide images included a retrospective cohort study that aimed to assess drug-related ovarian toxicity (22). The purpose of the study was to create a deep learning algorithm that is able to quantify corpora lutea (CLs) in H&E-stained ovarian tissue with similar accuracy to the pathologist’s assessment. The study was conducted on ovarian tissue of female Sprague-Dawley rats, and the sample was trained and validated using RetinaNet, the deep-learning algorithm of choice in this study. Due to the structural variability of rodent ovarian tissue, enumeration of CLs and evaluation of ovarian toxicity can be time consuming and challenging. Therefore, an automated method to quantify CLs would potentially decrease the workload for pathologists. Overall, drug-related ovarian toxicity was evident in the form of either increase or decrease in the number of CLs. In addition, mistakes in counting CLs by RetinaNet were similar to the pathologist’s variation, with a mean absolute difference of 0.57 + 0.096 between the pathologists and RetinaNet. Further training of the model with a further increase in sample size would improve the model’s accuracy (22).

A systematic review published in 2023 included a total final number of 45 studies, aiming to review the application of AI in pathological images for either diagnosis or prognosis of ovarian cancer (23). In these 45 studies, a total of 80 models were used including; convolutional neural network and its different architectures. CNN was the most commonly used (41/80), followed by support vector machine (SVM) (10/80), and random forest (6/80). Additionally, in most of the studies (18/45), the data source used for ovarian pathology slides is The Cancer Genome Atlas (TCGA), and other studies used their own data (12/45). Most researchers have used segmentation to determine the region of interest for tumor tissue. Most published research (nearly 37 papers) showed high or unclear risk of bias due to limited data used, such as; number of included patients and number of images (23). Another systematic review published in 2022 included 39 studies (4). Out of these studies, 7 used pathological images, 13 studies were based on serum tumor markers,

and the remaining 19 used high-throughput data. The review aimed to provide an objective assessment of AI algorithms application in the diagnosis and prediction of prognosis of ovarian cancer. The 7 studies that used pathological images to diagnose ovarian cancer showed specificity higher than 90%. However, there is a wide gap between the predictive performance of the current AI model and the clinicians’ experience, especially in prognostic prediction due to the narrow data set (4).

Herein, we present the preparation of a dataset of collected digital histopathological images of the major types of EOC, developing and training an AI model for automated identification of EOC subtypes, and evaluating its performance in comparison to pathologists’ diagnosis.

## 3 Methods

### 3.1 Dataset Preparation and Pre-processing

#### 3.1.1 Data Collection

Sixty-four public cases were retrieved from CIA (The Cancer Imaging Archive) including (20x) whole slide images (WSIs) and grouped into subtypes of epithelial ovarian cancer (clear cell carcinoma (CCC), endometrioid carcinoma (EC), mucinous carcinoma (MC) and serous carcinoma (SC)) (24).

In-house cases were collected from the archive of the pathology department at King Fahad University Hospital (KFUH) from 2014 to 2023, ending up with 18 Hematoxylin and Eosin (H&E) stained slides. The cases were classified into the four most common epithelial ovarian carcinoma subtypes (CCC, EC, MC, and SC). The diagnosis was reviewed and confirmed by experienced pathologists who assigned slide-level labels along with tumor annotation. The collected slides were scanned and digitized at 20x power by (I-Scan from Ventana), in our facility, to obtain 18 whole slide images in tiff format.

Both public and in-house cases were divided into training (80%) and evaluation (20%) subsets.

The Institutional Review Board (IRB) of Imam Abdulrahman Bin Faisal University approved the use of patients’ clinical information and histopathological samples (IRB number IRB-UGS-2023-01-410, approved on 22/10/2023).

##### Statement on Waiver of Consent

Consent for this study has been waived due to the retrospective nature of the research. The patients involved in the study are impossible to track. Additionally, the data utilized in this study were retrieved from archived microscopic slides and contained no patient identification, ensuring the anonymity of the subjects.

#### 3.1.2 Extract tiles from the WSIs

As the WSIs were huge (hundreds of thousands of pixels), it was impossible to process them as a whole. Hence fragmentation is crucial as the expected input size for the majority of deep learning models is 224 pixels. Therefore, all the public and in-house WSIs were cropped and resized into tiles of size (224 x 224 pixels). First, tumor regions of interest (ROIs) have been annotated by pathologists, then tiles were extracted from the ROIs, using QuPath software version 0.5 (25).

#### 3.1.3 Pre-processing Techniques

Considering the variation in color between the different WSIs, image normalization and color standardization are essential to normalize the obtained patches before introducing them to the model. For that purpose, we adopted the following Torchvision function: (transforms.Normalize(mean=[0.485, 0.456, 0.406], std=[0.229, 0.224, 0.225])).

### 3.2 Network Architectures and Training Pipeline

Considering the large size of the dataset, representative features from the resulting tiles were extracted and then used to train a classifier, rather than training the network directly on the images. This approach tremendously reduces the consumption of computing resources and the time for training the network.

#### 3.2.1 CNN Architectures

ResNet50, a CNN model, was used for features extraction. The model is pre-trained on ImageNet. It has been frozen after removing its classifier head to preserve pre-learned weight.

#### 3.2.2 Training Process

Two classifiers (CNN-based, and lightGBM) were selected for experimental training on the extracted features.

The CNN-based classifier was configured with specific parameters: Adam optimizer with a learning rate of 0.001 along with a scheduler, a batch size of 128, 21 epochs with early stopping, utilizing CrossEntropy loss function and Class-weighted-CrossEntropy. Additionally, cross-validation was incorporated during the training phase.

On the other hand, the lightGBM model was tuned with the following parameters: multiclass objective, a learning rate of 0.05, and multi_error metric. Early stopping was applied after 10 rounds, whereby training ceased if validation scores failed to improve for consecutive 10 rounds.

### 3.3 Performance Evaluation Metrics

- Accuracy: percentage of correct predictions
- Precision (specificity): accuracy of positive predictions.
- Recall (sensitivity): ability to identify all positive instances.
- F1-score: harmonic mean of precision and recall
- Confusion matrix for detailed analysis of classification errors

### 3.4 Experimental Setup

- Train-test split: 80% train, 20% test (stratified by class)
- Hardware: GPU (NVIDIA RTX3060 12GB), RAM 32GB, CPU i7 13^th^ gen.
- Implementation: PyTorch was used as Deep learning framework.

## 4 Results

### 4.1 Extracted tiles

Following the described methodology above, more than 2 million tiles/patches were extracted from the collected and annotated WSIs (public and in-house), using QuPATH (Table 1).

**Table 1:** Dataset details.

|  | CCC | EC | MC | SC | Total |
| --- | --- | --- | --- | --- | --- |
| Public WSIs | 25 | 5 | 6 | 28 | 64 |
| Public tiles | 750.000 | 90.000 | 110.000 | 850.000 | 1.800.000 |
| KFHU WSIs | 2 | 3 | 3 | 10 | 18 |
| KFHU tiles | 21.000 | 37.000 | 37.000 | 164.000 | 259.000 |

### 4.2 Networks prediction performance

Both networks (lightGBM and CNN) achieved comparable results with slightly higher performance for CNN (Table 2). However, the discriminative strength among classes was much better using Class-weighted CNN (Fig. 3).

**Table 2:** Evaluation metrics: Networks classification performance.

|  | Acc.<sup>1</sup> |  | Precision |  |  |  | Recall |  |  |  |  | F1 score |  |  |  |  |
| --- | --- | --- | --- | --- | --- | --- | --- | --- | --- | --- | --- | --- | --- | --- | --- | --- |
|  |  | Avg | CCC | EC | MC | SC | Avg | CCC | EC | MC | SC | Avg | CCC | EC | MC | SC |
| lightGBM | 0.97 | 0.95 | 0.98 | 0.91 | 0.94 | 0.97 | 0.91 | 0.99 | 0.79 | 0.9 | 0.99 | 0.93 | 0.98 | 0.84 | 0.92 | 0.98 |
| CNN | 0.98 | 0.96 | 0.99 | 0.9 | 0.96 | 0.99 | 0.95 | 0.99 | 0.9 | 0.95 | 0.99 | 0.95 | 0.99 | 0.9 | 0.95 | 0.99 |
| CNN (class-weighted) | 0.97 | 0.91 | 1 | 0.74 | 0.94 | 0.99 | 0.97 | 0.99 | 0.97 | 0.97 | 0.96 | 0.94 | 0.99 | 0.84 | 0.95 | 0.98 |

<sup>1</sup>Acc. : Accuracy

**Fig. 1:**
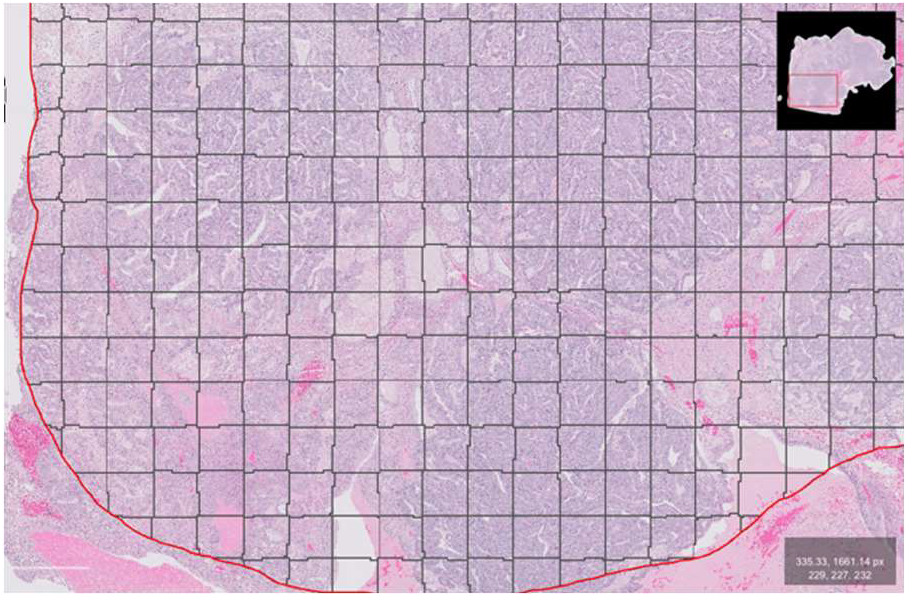
Tiles extracted from the annotated tumor region.

**Fig. 2:**
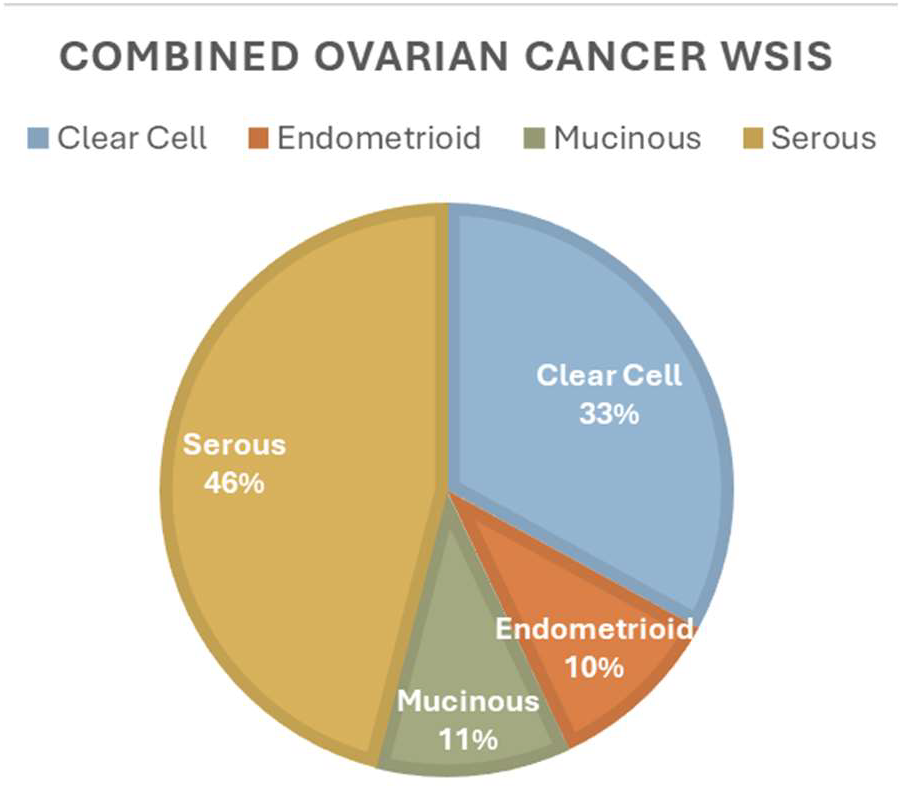
Distribution of the different classes in the combined dataset.

**Fig. 3:**
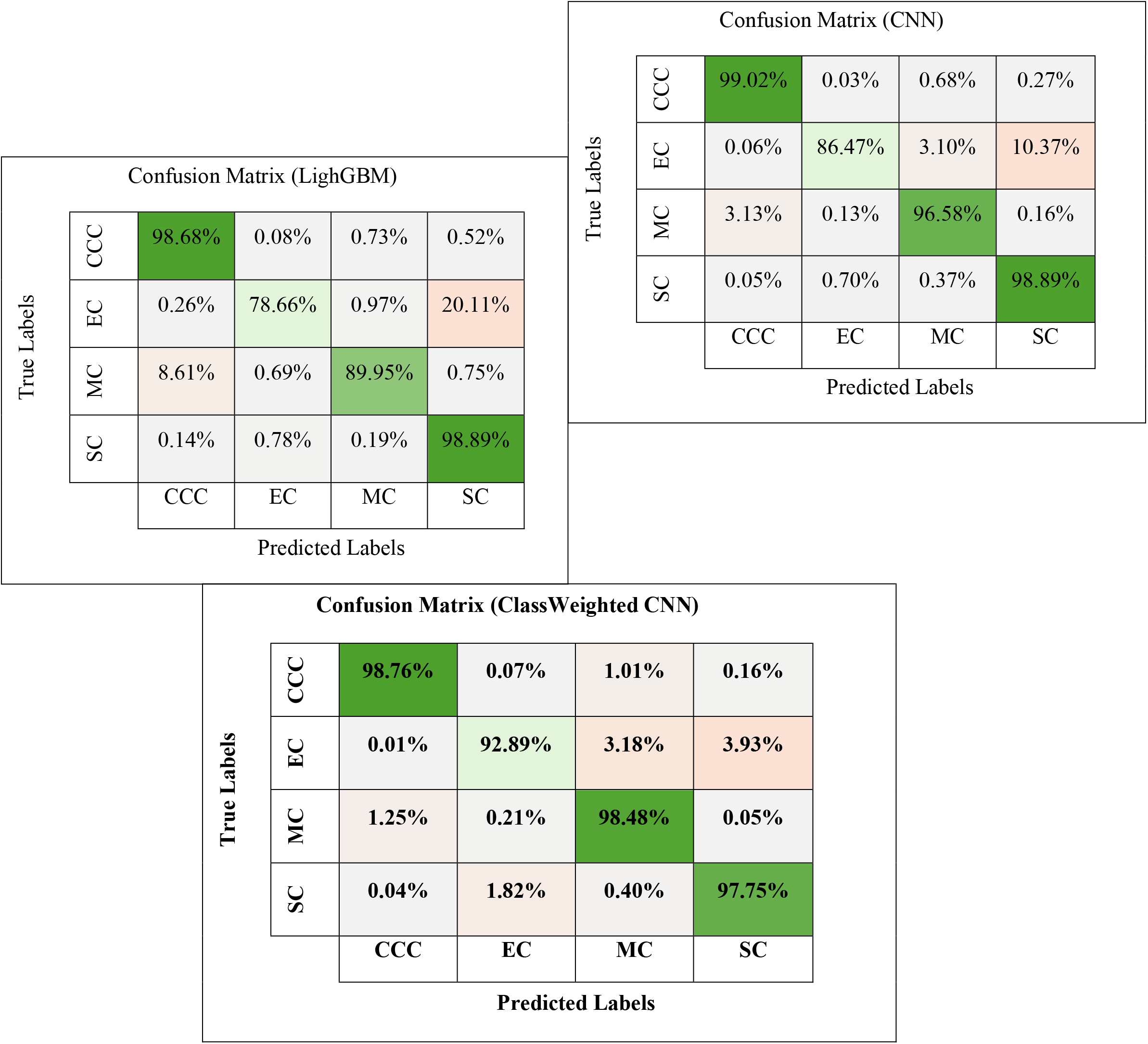
Detailed Comparison of the 3 classification networks performance (Percentage of correct predictions)

In our study the overall misclassification was in the following order from the highest to the lowest error rate: 1-EC misclassified as SC, 2-MC misclassified as CCC, 3-SC misclassified as EC, and 4-CCC misclassified as MC. Further elaboration about each classification model is demonstrated in Table 3.

**Table 3:** Misclassification error rate.

|  | LightGBM | CNN | CNN (class weighted) |
| --- | --- | --- | --- |
| EC misclassified as SC | 20.11% | 10.37% | 3.93% |
| MC misclassified as CCC | 8.61% | 3.13% | 1.25% |
| SC misclassified as EC | 0.78% | 0.70% | 1.82% |
| CCC misclassified as MC | 0.73% | 0.68% | 1.01% |

## 5 Discussion

Histopathology image analysis can be challenging and time-consuming for pathologists. An accurate diagnosis is essential to predict the prognosis, particularly with the different morphologic features and variations of each subtype of EOC. For this reason, the use of AI can provide a fast and accurate analysis of large and complex microscopic images. This study aimed to build an AI model that helps pathologists identify EOC subtypes incorporating local Saudi histopathological images. The AI classifiers developed in this study demonstrated promising performance in accurately predicting the major subtypes of epithelial ovarian cancer from digitized histopathology images. Utilizing deep learning techniques to extract discriminative features along with LightGBM and CNN classifiers resulted in patch-level accuracy rates of 97-98% on the test dataset. The CNN classifier showed particularly robust discriminative ability between the different cancer subtypes when class weighting was applied during training. These results align with previous studies highlighting the potential of AI models for ovarian cancer classification and diagnosis from histopathology images. The large and diverse dataset of over 2 million image tiles extracted from 82 whole slide images likely contributed to the strong generalization performance achieved by the models.

Although the overall accuracy is promising, F1 score of these network classifiers is variable among EOC subtypes. According to the F1 score, CCC shows a significantly higher score in lightGBM, CNN classifier, and CNN (class-weighted) with a score of (0.98,0.99,0.99), respectively, which is slightly similar to SC (0.98,0.99,0.98). Whereas EC scored the lowest (0.84,0.90,0.84). (Table 2).

EC is mostly misclassified as SC followed by MC misclassified as CCC, which is consistent with Wu et al findings. Wu et al denoted these misclassifications to the unclear morphology of some of the samples (16). According to Lim et al, 30% of OC were initially classified as EC and then reclassified to SC which emphasises the overlapping morphology between the two subtypes (26). As illustrated in Table 3, it is remarkable that all three models misclassify one specific subtype as another and vice versa (EC and SC \ MC and CCC) which strongly implies the intersecting morphological features rather than a model error.

However, it is important to note some limitations. First, a small in-house sample size. A large sample size is undoubtedly preferred to validate the AI model’s accuracy. Yet, KFUH has a limited data set, therefore, better accuracy and more generalizable results can be achieved by including more than one hospital or province. Second, subtypes sample number imbalance. Despite researcher efforts to assure all subtypes inclusion, overrepresentation of serous carcinoma was noticeable in this study sample as serous carcinoma is the most common EOC type in KSA and worldwide (6). Thus, the CNN class-weighted model was an attempt to overcome this limitation. Third, a shortage of computer resources restricts data input and analysis. Regardless of these limitations, the current paper provides an excellent accuracy level reaching up to 97-98% and encourages more work concerning AI application in general pathology settings and EOC classification.

The full end-to-end pipeline including the trained networks is very useful in conducting similar research in histopathology diagnoses and also could be used, after validation, as a helpful tool in daily cancer diagnosis. The full pipeline code will be available on https://github.com/hkussaibi.

## 6 Conclusion

This study provides evidence that AI-based computational pathology techniques can achieve human-level accuracy in ovarian cancer subtype classification from histopathology images. If properly validated, such AI-assisted diagnostic tools could greatly benefit pathology workflow by reducing subjectivity, decreasing inter-observer variability, and increasing efficiency compared to manual review alone. Future research should focus on prospective clinical validation and extend this approach to other cancer types and diagnostic tasks in digital pathology.

## Supporting information

IRB

## Data Availability

All data produced in the present study are available upon reasonable request to the authors

https://github.com/hkussaibi

