## Supplementary material for "Al-Powered classification of Ovarian cancers Based on Histopathological lmages": IRB

**Kingdom of Saudi Arabia**
**Ministry of Education**
**Imam Abdulrahman Bin Faisal University**
**Office of the Vice President for Research & Higher Studies**

جامعة الإمام عبدالرحمن بن فيصل
IMAM ABDULRAHMAN BIN FAISAL UNIVERSITY

المملكة العربية السعودية
وزارة التعليم
جامعة الإمام عبد الرحمن بن فيصل
وكالة الجامعة للدراسات
العليا والبحث العلمي

اللجنة الدائمة لأخلاقيات البحث على المخلوقات الحية
Institutional Review Board
NCBE Registration No.: (HAP-05-D-003)

| | | | |
|---|---|---|---|
| **IRB Number** | IRB-UGS-2023-01-410 | | أيرب-يوجس-١٠٤١-٠١-٢٠٢٣ |
| **Project Title** | AI-Powered classification of Ovarian Cancers Based on Histopathological Images | | |
| **Student Investigators** | Elaf Alibrahim, Eman Alamer, Shrooq Alshehab, Ghada Alhajji, Zahraa Shabib | | |
| **Supervisor** | Dr. Haitham Kussaibi | | |
| **College / Center** | Medicine | **Department** | Pathology |
| **Approval Date** | 22/10/2023 | | |

The application was reviewed and approved at Imam Abdulrahman Bin Faisal University IRB through an Expedited Review on Sunday, October 22, 2023.

Approval is given for one year from the date of approval. Projects, which have not commenced within six months of the original approval, must be re-submitted to the University Institutional Review Board (IRB) Committee. If you are unable to complete your research within the validation period, you will be required to request an extension from the IRB Committee.

On completion of the research, the Principal Investigator is required to advise the Institutional Review Board if any changes are made to the protocol, a revised protocol must be submitted to the Institutional Review Board for reconsideration.

Approval is given on the understanding that the "Guidelines for Ethical Research Practice" are adhered to. Where required, a signed written consent form must be obtained from each participant in the study group.

Chairman of the Institutional Review Board

Professor Badr Abdulrahman Aljandan

Stamp

25/10/2023

جامعة الإمام عبدالرحمن بن فيصل
IMAM ABDULRAHMAN BIN FAISAL UNIVERSITY
043 133 105

CC. - Dean
Deanship of Scientific Research
- Director General
King Fahd Hospital of the University
- Director
Center for Research and Medical Consultations
- Supervisor General for Quality and Safety
King Fahd Hospital of the University
- Director
Monitoring Office for Research and Research Ethics
-Director
Pharmacy @ KFHU
